# Loon Lens 1.0 Validation: Agentic AI for Title and Abstract Screening in Systematic Literature Reviews

**DOI:** 10.1101/2024.09.06.24313186

**Authors:** Ghayath Janoudi, Mara Uzun, Mia Jurdana, Ena Fuzul, Josip Ivkovic

## Abstract

**Introduction:** Systematic literature reviews (SLRs) are critical for informing clinical research and practice, but they are time-consuming and resource-intensive, particularly during Title and Abstract (TiAb) screening. Loon Lens, an autonomous, agentic AI platform, streamlines TiAb screening without the need for human reviewers to conduct any screening.

**Methods:** This study validates Loon Lens against human reviewer decisions across eight SLRs conducted by Canada’s Drug Agency, covering a range of drugs and eligibility criteria. A total of 3,796 citations were retrieved, with human reviewers identifying 287 (7.6%) for inclusion. Loon Lens autonomously screened the same citations based on the provided inclusion and exclusion criteria. Metrics such as accuracy, recall, precision, F1 score, specificity, and negative predictive value (NPV) were calculated. Bootstrapping was applied to compute 95% confidence intervals.

**Results:** Loon Lens achieved an accuracy of 95.5% (95% CI: 94.8–96.1), with recall at 98.95% (95% CI: 97.57–100%) and specificity at 95.24% (95% CI: 94.54–95.89%). Precision was lower at 62.97% (95% CI: 58.39–67.27%), suggesting that Loon Lens included more citations for full-text screening compared to human reviewers. The F1 score was 0.770 (95% CI: 0.734–0.802), indicating a strong balance between precision and recall.

**Conclusion:** Loon Lens demonstrates the ability to autonomously conduct TiAb screening with a substantial potential for reducing the time and cost associated with manual or semi-autonomous TiAb screening in SLRs. While improvements in precision are needed, the platform offers a scalable, autonomous solution for systematic reviews. Access to Loon Lens is available upon request at https://loonlens.com/.

## Introduction

Systematic literature reviews (SLRs) are essential for supporting clinical research, practice, and the development of health technologies.^1^ They are also increasingly becoming indispensable in fields such as psychology, social sciences, and computer sciences.^1^ The rigorous and methodical approach of SLRs ensures that critical decisions are based on the best available evidence, minimizing biases and providing a solid foundation for innovation and improved patient outcomes. However, conducting SLRs is extremely resource-intensive and time-consuming. The average SLR takes approximately 67.3 weeks to complete, with an estimated cost (as of 2019) of 141,194.80 USD.^2–4^

One of the most time-consuming steps in the SLR process is Title and Abstract (TiAb) screening, also known as level-1 (L1) screening. Dual screening, the recommended method, can require up to 1,089 person-hours and cost approximately 75,139 GBP on a project with 12,477 abstracts.^5,6^ Automating this labor-intensive process has thus been an active area of research.^7,8^ However, several challenges hinder effective automation from a machine learning perspective—most notably, class imbalance and generalizability.

Class imbalance is a significant issue in TiAb screening, where up to 95% of the initially identified citations can be excluded after screening.^2,9^ Generalizability, on the other hand, refers to the difficulty of applying a model trained on one SLR to another SLR. This is because SLRs can vary significantly in scope, topic, and purpose.^1^ These challenges have given rise to semi-automated methods, often using active learning.^10,11^ In active learning approaches, a human reviewer labels a subset of records, which are then used to train a machine learning model tailored to the specific SLR.^11^ This method has been estimated to reduce the workload by half, translating into a cost of around 37,860 GBP for a project of 12,477 abstracts.^5,6,12^

Recently, large language models (LLMs) have emerged as a promising new avenue for tackling TiAb screening in an entirely autonomous manner, without human involvement. While early attempts at using LLMs for TiAb screening have shown promise, concerns remain about scalability, usability, reproducibility, and the extent of validation conducted.^13–16^ There is therefore a need for a validated, scalable, intuitive, and specialized tool that can conduct TiAb screening autonomously, without requiring pre-labeled data.

Loon Lens is an LLM-based agentic AI platform designed to address these needs. It conducts unsupervised TiAb screening in a scalable and intuitive way, requiring only the inclusion and exclusion criteria to perform the screening. The user needs to upload their citations to the platform in the form of a RIS file, input their inclusion and exclusion criteria, and click on a button for Loon Lens to screen all of the citations. In this paper, we conduct a validation study and report the results of the diagnostic accuracy of Loon Lens.

## Methods

The reporting of this validation study adhered to the reporting guidelines outlined in STARD, where applicable.^17^

### Study Design

This is a prospective validation study with the primary objective of evaluating the performance of Loon Lens, an agentic AI platform for autonomous Title and Abstract (TiAb) screening, against ground truth decisions derived from human reviewers. A total of eight SLRs were replicated to produce the validation dataset. These SLRs were originally conducted by Canada’s Drug Agency (CDA) to inform drug reimbursement decisions. These eight SLRs represented a sample of convenience of the recent available reviews where the eligibility criteria (inclusion and exclusion criteria) are clearly outlined in the report. The included SLRs were conducted to provide evidence on the following drugs: Darolutamide (brand name Nubeqa),^18^ Durvalumab (brand name Imfinzi),^19^ Crisantaspase Recombinant (brand name Rylaze),^20^ Upadacitinib (brand name Rinvoq),^21^ Guselkumab (brand name Tremfya),^22^ Lumasiran (brand name Oxlumo),^23^ Mepolizumab (brand name Nucala),^24^ Finerenone (brand name Kerendia).^25^.

### Validation Data Collection and Labeling

#### Search Strategy

For each systematic review, we developed a search strategy based on the search terms reported in the CDA reimbursement reports. The searches were conducted using OpenAlex, an open-source scholarly database.^26^ The search strategy for each SLR is presented in Appendix A – Search strategies. Searches were updated frequently throughout the conduct of the validation study with the last update being on September 1, 2024.

#### Eligibility Criteria

The eligibility criteria used in the CDA clinical reports for reimbursement were applied directly with minor modifications. These adjustments were primarily related to ensuring that the pivotal trials that were included by default in CDA SLRs also match the eligibility criteria. The eligibility criteria for each SLR are outlined in Appendix B – Inclusion Criteria.

#### Title and Abstract Screening

Two independent reviewers performed TiAb screening for all retrieved citations (GJ, MU, MJ, EF, JI). Disagreements between the reviewers were resolved through adjudication with another reviewer (GJ or MU). The agreed-upon decisions from the dual reviewers or the decision from the adjudicator served as the ground truth for evaluating the performance of Loon Lens.

### Test Methods

After establishing the ground truth for each systematic review, the same citations were screened using Loon Lens. Loon Lens is an LLM-based agentic AI platform designed to perform unsupervised screening of TiAb based solely on provided inclusion and exclusion criteria. The platform autonomously screens the titles and abstracts, and its predictions were compared to the ground truth labels.

#### Metrics Calculation

To assess the diagnostic performance of Loon Lens in TiAb screening, we computed several metrics commonly used in binary classification:

- **Accuracy**: The proportion of correct predictions (both positive and negative) made by Loon Lens.
- **Recall (Sensitivity)**: The proportion of actual positives correctly identified by Loon Lens.
- **Precision (Positive Predictive Value - PPV)**: The ratio of correctly identified positive instances to all instances identified as positive by Loon Lens.
- **F1-score**: The harmonic mean of precision and recall.
- **F-beta score (with beta = 10)**: A weighted metric that prioritizes recall over precision.
- **Specificity**: The proportion of actual negatives that were correctly identified by Loon Lens.
- **Negative Predictive Value (NPV)**: The ratio of correctly identified negative instances to all instances predicted as negative by the system.

A confusion matrix was generated to depict the counts of true positives (TP), true negatives (TN), false positives (FP), and false negatives (FN), which formed the basis for the calculation of these metrics.

#### Bootstrapping for Confidence Intervals

To assess the uncertainty of the performance metrics, we applied bootstrapping. The bootstrapping method involved resampling the dataset with replacement 1,000 times to compute 95% confidence intervals (CI) for each metric. Each iteration consisted of the following:

1. **Resampling**: Both the ground truth labels and the predictions made by Loon Lens were resampled with replacement.
2. **Metric Calculation**: For each resample, we recalculated the following metrics: accuracy, precision (PPV), recall (sensitivity), F1-score, specificity, NPV, and F-beta score.
3. **Confidence Interval Estimation**: The 95% CIs for each metric were computed using the percentile method, taking the 2.5th and 97.5th percentiles from the distribution of the bootstrapped results.

#### Software and Tools

All analyses were performed using Python.^27^ The scikit-learn library was used for metric computation and resampling, while pandas was employed for data handling.^28,29^ The numpy package facilitated the bootstrapping procedure.^30^

#### Access to Loon Lens

Access to the Loon Lens platform for TiAb screening is available at the following URL: https://loonlens.com/.

## Results

### Validation Data

For the 8 included SLRs, we retrieved a total of 3,796 citations from OpenAlex bibliographic database. Of these, the human screening process identified a total of 287 (7.6%) citations as ‘included’. A breakdown of the number of citations and the determination of the human screeners per SLR is presented in Table 1.

**Table 1:** Number of citations and total included per human reviewers.

| SLR | Total citations, N | Included – human reviewers, n (%) |
| --- | --- | --- |
| PC0294 Nubeqa | 403 | 31 (7.7) |
| PC0296 Imfinzi | 295 | 9 (3.1) |
| PC0301 Rylaze | 1,365 | 10 (0.7) |
| SR0730 Rinvoq | 369 | 52 (14.1) |
| SR0733 Tremfya | 679 | 82 (12.1) |
| SR0734 Oxlumo | 148 | 24 (16.2) |
| SR0735 Nucala | 172 | 17 (9.9) |
| SR0737-Kerendia | 365 | 62 (17.0) |
| **Total** | **3,796** | **287 (7.6)** |

## Test Results

The diagnostic performance of Loon Lens was assessed against a ground truth determined by human reviewers’ screening decisions. Several performance metrics were calculated, including accuracy, precision, recall, F1 score, specificity, F-beta score, NPV, and PPV. These metrics, along with their 95% confidence intervals (CI), are summarized in Table 2. The diagnostic performance results are also presented in In addition, the performance of Loon Lens in classifying the citations is detailed in the confusion matrix Table 3, which compares the predicted outcomes by Loon Lens against the actual classification by the human reviewers.

**Table 2:** Performance metrics of Loon Lens in TiAb screening.

| Metric | Value | 95% CI |
| --- | --- | --- |
| Accuracy | 0.955 | 0.948 – 0.961 |
| Recall (Sensitivity) | 0.990 | 0.976 – 1.000 |
| Precision (PPV) | 0.630 | 0.584 – 0.673 |
| F1 Score | 0.770 | 0.734 – 0.802 |
| **Specificity** | 0.952 | 0.945 – 0.959 |
| **F-beta Score** | 0.984 | 0.970 – 0.994 |
| **NPV** | 0.999 | 0.998 – 1.000 |

**Table 3:**
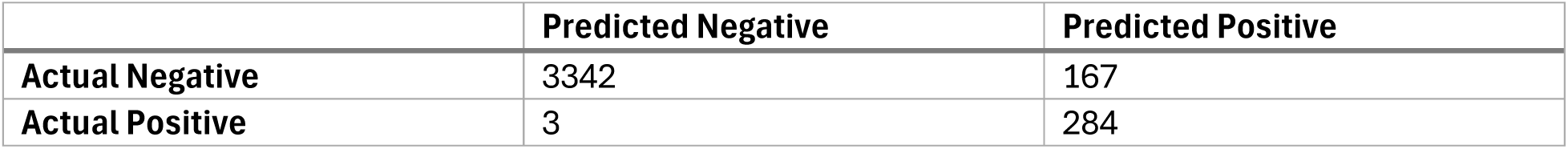
Confusion Matrix.

**Figure 1:**
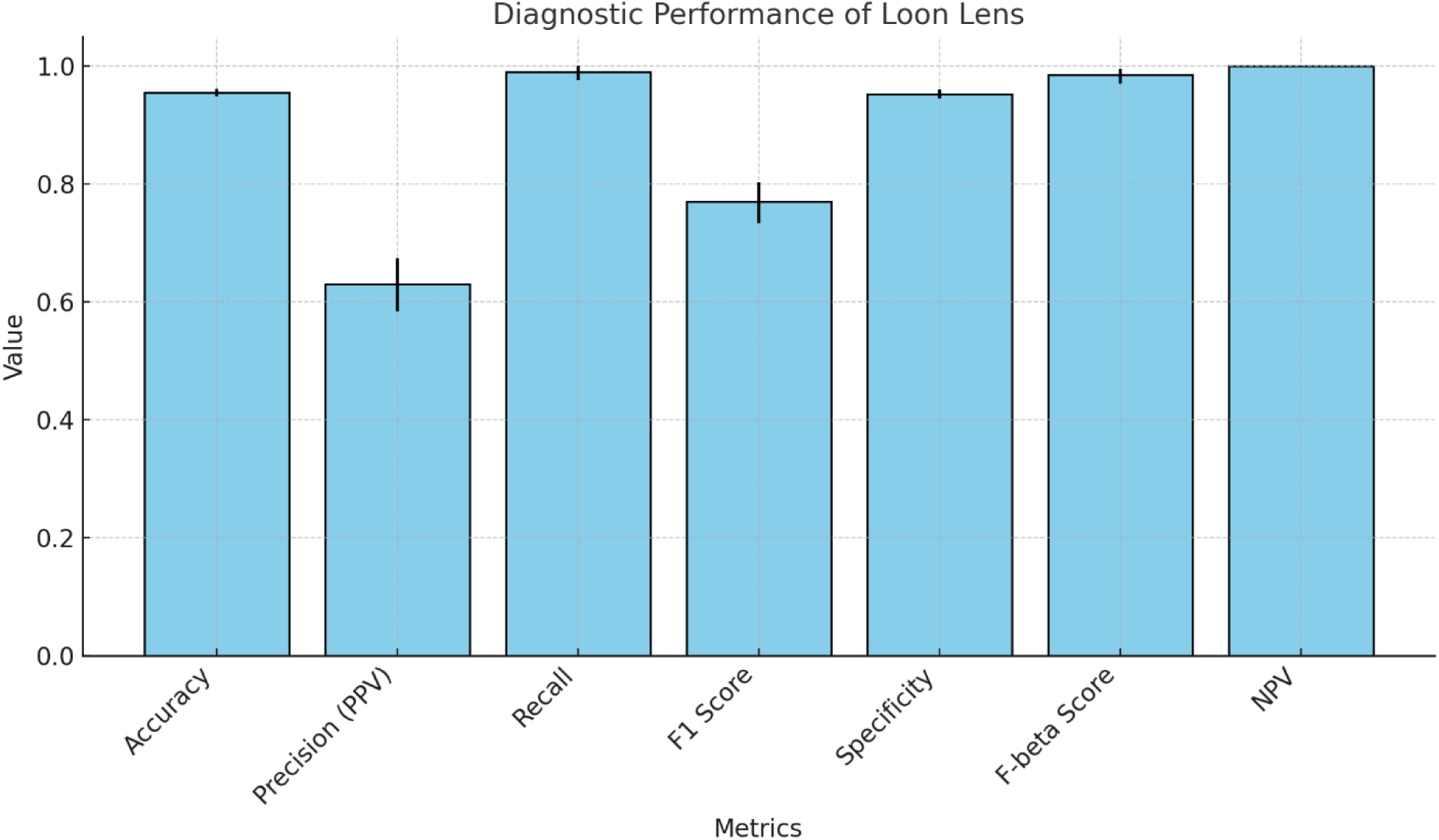
Diagnostic Performance of Loon Lens

Loon Lens demonstrated strong overall performance in accurately classifying citations. It achieved an accuracy of 95.5% (95% CI: 94.8%–96.1%), indicating that the model correctly classified the majority of citations. The recall, or sensitivity, was very high at 98.95% (95% CI: 97.57%–100%), indicating that Loon Lens successfully identified almost all citations that should have been included. Specificity, the ability to correctly exclude irrelevant citations, was 95.24% (95% CI: 94.54%–95.89%). Precision was 62.97% (95% CI: 58.39%–67.27%), meaning that around 63% of the citations flagged as ‘included’ by Loon Lens were indeed correctly classified as included by human screeners.

The F1 score, which balances precision and recall, was 0.770 (95% CI: 0.734–0.802), highlighting the strong performance of the model in this screening task. The confusion matrix reveals that out of the 3,345 actual positives, the platform only misclassified 3 citations as negatives. Conversely, it misclassified 167 of the 3,342 actual negatives as positives.

## Discussion

The primary aim of this study was to evaluate the diagnostic performance of Loon Lens, an LLM-based agentic AI platform, for fully autonomous TiAb (or level 1) screening in SLRs. Our findings indicate that Loon Lens demonstrated strong overall performance, with high recall (sensitivity) and specificity rates, making it a promising tool for automating this labor-intensive process.

As SLRs aims to capture all of the evidence that addresses the research question, any missing relevant studies could undermine the overall conclusions. Thus, it is essential for any classifier to achieve high recall (sensitivity). With a recall of 98.95%, Loon Lens nearly matched human performance, successfully identifying almost all relevant citations.

However, recall (sensitivity) alone is not a sufficient indication of the usefulness of a model. A model that labels all the citations as ‘included’ would achieve a recall of 100%. But such a model would not be useful to automate the task of screening. Specificity and precision play an important role in reflecting upon the model ability to exclude citations and include citations that may not need to be included. The specificity of 95.24% shows that Loon Lens effectively excluded irrelevant studies, minimizing the number of false positives. The precision of 62.97% indicates that about 37% of the studies classified as relevant by Loon Lens were actually false positives. This would suggest that Loon Lens may send more articles for full-text screening than a human reviewer would. Considering the large benefits in automating this task, an additional 37% of full-text screening is considered acceptable. In addition, expanding Loon Lens to cover the full-text screening is likely to reduce the impact of false positives.

The F1 score of 0.770 further confirms the strong balance between precision and recall. Given that the F1 score is the harmonic mean of these two metrics, it highlights Loon Lens’s ability to maintain a relatively low false-negative rate while keeping the number of false positives manageable. Additionally, the high F-beta score (0.984), which emphasizes recall over precision, is particularly relevant in contexts where false negatives (i.e., missed relevant studies) are more costly than false positives.

Our findings align with previous studies exploring the use of AI in TiAb screening, where semi-automated and machine learning methods have shown the potential to reduce the manual workload by as much as 50%.^5,6,12^ However, Loon Lens’s fully autonomous approach sets it apart from prior models that rely heavily on active learning or pre-labeled training data. This validation study demonstrates that Loon Lens can operate without prior human review to label citations during screening, presenting a significant leap forward in scalability and efficiency.

This validation study tested Loon Lens across eight different SLRs with varying subject areas and inclusion/exclusion. These results indicate its ability to generalize beyond the specific scope of training datasets.

One of the key strengths of this study is the use of real-world, high-stakes SLRs in the healthcare domain, particularly drug reimbursement decisions. These SLRs involve critical clinical evidence, practice forming decisions and recommendations that would determine patient access to new and innovative therapeutics. Thus, the ability of Loon Lens to perform at a high level in this context underscores its practical applicability.

However, there are limitations that should be acknowledged. First, the study included only eight SLRs, which, while diverse in topic, may not capture the full range of complexity found in SLRs across other fields like psychology or social sciences. Furthermore, all of the included SLRs were of interventional studies. Thus, the presented results may not be directly generalizable to other type of SLRs that focus on different designs. Future research should aim to replicate this study with a broader set of SLRs across different disciplines and designs to confirm generalizability. Second, while Loon Lens demonstrated high recall and specificity, the lower precision suggests that human intervention may still be required to confirm the citations flagged for inclusion, particularly in cases where accuracy is critical.

Another limitation relates to the data source. The use of OpenAlex as the bibliographic database may introduce biases depending on the scope and completeness of the database. While it is an open-source and comprehensive tool, comparisons with other databases such as PubMed or Scopus may yield different results. Furthermore, while bootstrapping provided robust confidence intervals for the performance metrics, future studies should explore alternative methods for uncertainty quantification, especially in edge cases where AI models may struggle.

There are several areas for future improvement and research. One key avenue is refining the model’s precision to reduce false positives. This can be achieved through further fine-tuning of the Loon Lens algorithm. Additionally, future work should focus on expanding the validation of Loon Lens across a wider range of disciplines and document types (e.g., conference papers, grey literature). Exploring its performance in different SLR contexts, such as those with a higher proportion of qualitative studies or non-RCTs, would help assess the platform’s versatility.

## Conclusion

Loon Lens has demonstrated high diagnostic accuracy, recall (sensitivity), and specificity in autonomous TiAb screening for systematic literature reviews. Its ability to generalize across multiple SLR topics and deliver performance comparable to human reviewers highlights its potential to significantly reduce the time and cost associated with manual screening. While there is room for improvements in precision, the overall balance between fully autonomous TiAb screening and the additional cost of false positives must be taken into account. In conclusion, Loon Lens presents an immediately applicable and promising solution for automating a traditionally resource-intensive process.

## Appendix A Search strategies

### PC0294 Nubeqa

((darolutamide OR “darolutamide”) OR (nubeqa OR “nubeqa”) OR (“darramamide” OR darramamide) OR (“bay-1841788” OR “bay1841788” OR “odm-201” OR “odm201” OR “orm-16497” OR “orm16497” OR “orm-16555” OR “orm16555” OR “X05U0N2RCO”)) AND (((randomized OR randomised) AND (trial OR study OR research OR clinical OR controlled)) OR (phase AND (I OR II OR III OR IV OR 1 OR 2 OR 3 OR 4 OR one OR two OR three OR four)) OR “placebo” OR “double-blind” OR “double_blind” OR “double blind” OR “open label” OR “open-label” OR “open_label” OR “single-blind” OR “single_blind” OR “single blind”)

### PC0296 Imfinzi

(((((bile OR hepatic OR gall OR gallbladder OR (gall AND bladder)) AND (duct OR tract)) OR hepatobiliary OR hepatocellular OR choledochus OR colangio OR ampulla) AND (cancer OR tumor OR tumour OR carcinoma OR neoplasm OR malignant OR adenocarcinoma)) OR cholangiocarcinoma) AND ((durvalumab OR “durvalumab”) OR (imfinzi OR “imfinzi”) OR (“medi 4736” OR “medi4736”)) AND (((randomized OR randomised) AND (trial OR study OR research OR clinical OR controlled)) OR (phase AND (I OR II OR III OR IV OR 1 OR 2 OR 3 OR 4 OR one OR two OR three OR four)) OR “placebo” OR “double-blind” OR “double_blind” OR “double blind” OR “open label” OR “open-label” OR “open_label” OR “single-blind” OR “single_blind” OR “single blind”)

### PC0301 Rylaze

((“Asparaginase” OR Asparaginase) OR (“rylaze” OR rylaze) OR (“crisantaspas” OR crisantaspas) OR (“erwinase” OR erwinase) OR (“erwinaze” OR erwinaze) OR (“jzp 458” OR “jzp458” OR “D733ET3F9O”)) AND (((randomized OR randomised) AND (trial OR study OR research OR clinical OR controlled)) OR (phase AND (I OR II OR III OR IV OR 1 OR 2 OR 3 OR 4 OR one OR two OR three OR four)) OR “placebo” OR “double-blind” OR “double_blind” OR “double blind” OR “open label” OR “open-label” OR “open_label” OR “single-blind” OR “single_blind” OR “single blind”)

### SR0730 Rinvoq

((ulcerative AND colitis) OR (“ulcerative colitis”) OR (Colitis OR “colitis”) OR (proctocolitis OR “proctocolitis”) OR (colorectitis OR “colorectitis”)) AND ((“rinvoq” OR rinvoq) OR (“upadacitinib” OR upadacitinib) OR (“abt494” OR “abt 494” OR “NEW4DV02U5”))

### SR0733 Tremfya

((tremfya OR “tremfya”)OR (guselkumab OR “guselkumab”) OR (“cnto 1959” OR “cnto1959” OR “089658A12D”)) AND (((randomized OR randomised) AND (trial OR study OR research OR clinical OR controlled)) OR (phase AND (I OR II OR III OR IV OR 1 OR 2 OR 3 OR 4 OR one OR two OR three OR four)) OR “placebo” OR “double-blind” OR “double_blind” OR “double blind” OR “open label” OR “open-label” OR “open_label” OR “single-blind” OR “single_blind” OR “single blind”)

### SR0734 Oxlumo

((lumasiran OR “lumasiran”) OR (Oxlumo OR “Oxlumo”) OR (“ad 65585” OR “ad65585” OR “aln 65585” OR “aln65585” OR “aln g01” OR “alng01” OR “aln go1” OR “alngo1” OR “RZT8C352O1” OR “67P6XH37HD”))

### SR0735 Nucala

((((rhino AND sinusitis) OR (rhinosinusitis) OR (sinus AND inflammation) OR Sinusitis OR rhinitis) AND (chronic OR persistent OR recurrent OR flareup OR “flare up”)) OR ((nasal AND polyp) OR rhinopolyp OR “CRSwNP”)) AND ((Nucala OR “Nucala”) OR (mepolizumab OR “mepolizumab”) OR (bosatria OR “bosatria”) OR (“90Z2UF0E52” OR “SB240563” OR “SB 240563”)) AND (((randomized OR randomised) AND (trial OR study OR research OR clinical OR controlled)) OR (phase AND (I OR II OR III OR IV OR 1 OR 2 OR 3 OR 4 OR one OR two OR three OR four)) OR “placebo” OR “double-blind” OR “double_blind” OR “double blind” OR “open label” OR “open-label” OR “open_label” OR “single-blind” OR “single_blind” OR “single blind”)

### SR0737-Kerendia

((finerenone OR “finerenone”) OR (kerendia OR “kerendia”) OR (“BAY94-8862” OR “BAY-94-8862” OR “BAY948862” OR “BAY-948862” OR “DE2O63YV8R”)) AND (((randomized OR randomised) AND (trial OR study OR research OR clinical OR controlled)) OR (phase AND (I OR II OR III OR IV OR 1 OR 2 OR 3 OR 4 OR one OR two OR three OR four)) OR “placebo” OR “double-blind” OR “double_blind” OR “double blind” OR “open label” OR “open-label” OR “open_label” OR “single-blind” OR “single_blind” OR “single blind”)

## Appendix B Inclusion Criteria

### PC0294 Nubeqa

**Population**

– Patients with metastatic castration-sensitive prostate cancer who are chemotherapy-eligible.
– Subgroups:
– ECOG performance status (Eastern Cooperative Oncology Group)
– Gleason score
– Extent of metastatic disease (e.g., lymph nodes, bone, viscera)
– Prior ADT (androgen deprivation therapy)
– Prior docetaxel therapy

**Intervention**

– Darolutamide 600 mg twice daily orally + docetaxel + ADT.

**Comparators**

– Apalutamide + ADT
– Enzalutamide + ADT
– Docetaxel + ADT
– Abiraterone + ADT + prednisone
– Abiraterone + ADT + prednisone + docetaxel
– Placebo

**Outcomes**

– Efficacy outcomes:
– OS (overall survival)
– Time to castration-resistant prostate cancer
– Time to initiation of subsequent antineoplastic therapy
– Time to pain progression
– HRQoL (health-related quality of life)
– ORR (objective response rate)
– Time to skeletal-related events
– Time to PSA (prostate-specific antigen) progression
– PSA response rates
– Harms outcomes:
– AEs (adverse events)
– SAEs (serious adverse events)
– WDAEs (withdrawal due to adverse events)
– Mortality

**Study Design**

– Published and unpublished phase III and IV RCTs (randomized controlled trials)

### PC0296 Imfinzi

**Population**

– Patients with locally advanced or metastatic BTC (biliary tract cancer).
– Subgroups:
– Sex (male vs. female)
– Disease status (locally advanced vs. metastatic)
– Primary tumour location (IHCC [intrahepatic cholangiocarcinoma] vs. EHCC [extrahepatic cholangiocarcinoma] vs. GBC [gall bladder cancer])
– PD-L1 status (programmed cell death 1 ligand 1)
– ECOG PS (Eastern Cooperative Oncology Group Performance Status)
– Geographic region

**Intervention**

– Durvalumab 1,500 mg IV (intravenous) plus chemotherapy every 3 weeks, followed by 1,500 mg IV every 4 weeks as monotherapy.

**Comparator**

– Gemcitabine alone or in combination with cisplatin or carboplatin.
– Placebo

**Outcomes**

– Efficacy outcomes:
– OS (overall survival)
– PFS (progression-free survival)
– Clinical response (i.e., ORR [objective response rate], DOR [duration of response], DCR [disease control rate])
– TTD (time to treatment discontinuation)
– HRQoL (health-related quality of life)
– Symptom severity (e.g., PGI-S [Patient Global Impression of Severity])
– Treatment tolerability
– Biochemical tumour markers of response
– Harms outcomes:
– AEs (adverse events)
– SAEs (serious adverse events)
– WDAEs (withdrawal due to adverse events)
– Mortality
– Notable harms/harms of special interest:
– Immune-mediated AEs (e.g., pneumonitis)
– Infusion-related reactions
– Infections (e.g., cholangitis, biliary tract infections)
– GI events (e.g., diarrhea)

**Study Designs**

– Published and unpublished phase III and IV RCTs (randomized controlled trials)

### PC0301 Rylaze

**Patient Population**

– Adult and pediatric populations aged 1 year and older receiving a multidrug chemotherapeutic regimen for the treatment of ALL (acute lymphoblastic leukemia) and LBL (lymphoblastic lymphoma) who have developed hypersensitivity to E. coli–derived asparaginase.
– Subgroups:
– Age group (e.g., < 25 years vs. ≥ 25 years)
– ALL vs. LBL
– CNS (central nervous system) involvement

**Intervention**

– Crisantaspase recombinant 25 mg/m² on Monday and Wednesday and 50 mg/m² on Friday, administered intramuscularly, for a total of 6 doses (to replace each planned dose of long-acting E. coli–derived asparaginase).

**Comparators**

– Erwinia-derived asparaginase
– No comparator

**Outcomes**

– Efficacy outcomes:
– Overall survival
– Event-free survival
– Disease-free survival
– Complete clinical remission and/or minimal residual disease
– NSAA (nadir serum asparaginase activity) levels
– HRQoL (health-related quality of life)
– Harms outcomes:
– AEs (adverse events), SAEs (serious adverse events), and WDAEs (withdrawal due to adverse events)
– Mortality
– Notable harms: thrombosis, pancreatitis, hemorrhage, hypersensitivity reaction, and hepatotoxicity

**Study Design**

– Phase II and Phase III RCTs (randomized controlled trials) and single arm trials

### SR0730 Rinvoq

**Population**

– Adult patients with moderately to severely active ulcerative colitis who have had an inadequate response, loss of response, or were intolerant to either conventional therapy or a biologic drug (i.e., TNF alpha antagonists, integrin receptor antagonists, or interleukin 12 or interleukin 23 inhibitors).
– Subgroups:
– Patients with previous vs. no previous conventional therapy
– Patients with previous vs. no previous biologic therapy
– Disease severity (moderate vs. severe)
– Disease extent (extensive vs. limited colitis)
– Primary nonresponders vs. secondary loss of response

**Intervention**

– Upadacitinib, oral tablets.
– Induction: 45 mg once daily for 8 weeks; an additional 8 weeks of 45 mg once daily may be needed for patients who do not achieve adequate therapeutic benefit by week 8.
– Maintenance: 15 mg or 30 mg once daily.

**Comparator**

– Adalimumab
– Golimumab
– Infliximab
– Tofacitinib
– Ustekinumab
– Vedolizumab
– Conventional therapy (i.e., any combination of aminosalicylates, corticosteroids, and/or immunomodulators)
– Placebo

**Outcomes**

– Efficacy outcomes:
– Clinical remission (including corticosteroid-free clinical remission)
– Clinical response
– Endoscopic remission
– Endoscopic improvement
– Histologic remission
– Histologic improvement
– Mucosal healing
– Symptoms relief (e.g., abdominal pain, rectal bleeding, bowel urgency)
– Health-related quality of life
– Need for colectomy
– Extraintestinal manifestations (e.g., fever, inflammation of the eyes or joints, mouth or skin ulcers, tender and inflamed nodules on shins)
– Emergency department visits or hospitalization
– Work productivity
– Harms outcomes:
– AEs (adverse events), SAEs (serious adverse events), WDAEs (withdrawal due to adverse events), mortality
– Notable harms (e.g., serious or opportunistic infection, malignancy, thrombosis, hypersensitivity, hepatotoxicity, anemia, lymphopenia, neutropenia, gastrointestinal perforation, hyperlipidemia)

**Study Designs**

– Published and unpublished phase III and IV RCTs (randomized controlled trials)

### SR0733 Tremfya

**Patient Population**

– Adults with active psoriatic arthritis.
– Subgroups:
– Previous exposure to bDMARDs (biologic disease-modifying antirheumatic drugs) (e.g., treatment-naive; treatment-experienced; non-responsive or intolerant)
– Concomitant treatment with non-biologic DMARDs

**Intervention**

– Guselkumab 100 mg SC (subcutaneous) injection at week 0 and week 4, followed by maintenance dosing every 8 weeks thereafter (alone or in combination with a cDMARD [e.g., methotrexate]).

**Comparators**

– TNF inhibitors (adalimumab, certolizumab pegol, etanercept, golimumab, infliximab)
– IL-17 inhibitors (ixekizumab, secukinumab)
– IL-12/23 inhibitors (ustekinumab)
– IL-23 inhibitors (risankizumab)
– JAK inhibitors (tofacitinib, upadacitinib)
– Apremilast
– Abatacept
– cDMARDs (e.g., methotrexate) alone or in combination with biologic or other DMARDs
– Placebo

**Outcomes**

– Efficacy outcomes:
– Clinical response in PsA (psoriatic arthritis) symptoms (e.g., ACR 20, ACR 50, ACR 70, MDA, DAS 28)
– Measure of function and disability (e.g., HAQ-DI)
– Health-related quality of life
– Measure of skin disease (e.g., PASI 75, PASI 90, PASI 100, or IGA response)
– Measure of other musculoskeletal disease (e.g., dactylitis, enthesitis, and axial arthritis)
– Measure of PsA symptoms (e.g., pain, fatigue)
– Radiologic changes
– Harms outcomes:
– AEs (adverse events), SAEs (serious adverse events), WDAEs (withdrawal due to adverse events), mortality, and notable harms (serious infections, hypersensitivity reactions, elevated hepatic enzymes, hepatic disorders, injection-site reactions)

**Study Design**

– Published and unpublished phase III and IV RCTs (randomized controlled trials)

### SR0734 Oxlumo

**Population**

– Pediatric and adult patients with primary hyperoxaluria type 1.
– Subgroups:
– Age
– Kidney function (e.g., eGFR)
– Baseline urinary and/or plasma oxalate levels
– Genetic status (e.g., G170R homozygous vs. G170R heterozygous vs. other)

**Intervention**

– Lumasiran administered via subcutaneous injection with weight-based loading and maintenance dosing:
– < 10 kg: Loading dose of 6 mg/kg monthly for 3 doses and maintenance dose of 3 mg/kg monthly
– 10 kg to < 20 kg: Loading dose of 6 mg/kg monthly for 3 doses and maintenance dose of 6 mg/kg once every 3 months (quarterly)
– ≥ 20 kg: Loading dose of 3 mg/kg monthly for 3 doses and maintenance dose of 3 mg/kg once every 3 months (quarterly)

**Comparator**

– Standard of care (e.g., dietary changes, hyperhydration, citrate supplementation, vitamin B6, dialysis, liver-kidney transplant)
– Placebo
– No comparator

**Outcomes**

– Efficacy outcomes:
– Kidney function (eGFR or creatinine levels)
– Loss of kidney function over time
– Prevention of dialysis and/or liver-kidney transplant
– Kidney stone events (e.g., severity)
– HRQoL (health-related quality of life)
– Urinary oxalate levels
– Plasma oxalate levels
– Urine oxalate:creatinine measures
– Harms outcomes:
– AEs (adverse events)
– SAEs (serious adverse events)
– WDAEs (withdrawal due to adverse events)
– Mortality
– Notable harms and harms of special interest:
– Injection site reactions
– Renal events
– Complications from systemic oxalosis
– Headache
– Rhinitis
– Upper respiratory infection
– Hypersensitivity reactions
– ADAs (antidrug antibodies)

**Study Designs**

– Published and unpublished phase III and IV RCTs (randomized controlled trials) and single-arm clinical trials

### SR0735 Nucala

**Population**

– Adult patients 18 years and older with severe CRSwNP (chronic rhinosinusitis with nasal polyps) inadequately controlled by intranasal corticosteroids alone.
– Subgroups:
– Asthma diagnosis (yes/no)
– Prior surgery (yes/no)

**Intervention**

– Mepolizumab, 100 mg administered by SC (subcutaneous) injection once every 4 weeks, used in combination with intranasal corticosteroids and/or saline irrigation.

**Comparator**

– Intranasal corticosteroids and/or saline irrigation.
– Placebo

**Outcomes**

– Efficacy outcomes:
– Nasal obstruction
– VAS (visual analogue scale) for nasal obstruction
– Symptoms:
– Composite VAS symptom score for nasal discharge, feeling of mucus in the throat, loss of smell, facial pain, and nasal polyp symptoms
– Sense of smell
– Response to treatment:
– Change in nasal polyp size
– Severity of nasal polyps and nasal obstruction:
– Endoscopic nasal polyp score
– Nasal congestion:
– PnIF (peak nasal inspiratory flow)
– HRQoL (health-related quality of life):
– SNOT-22 (Sino-Nasal Outcome Test 22)
– Systemic steroid use for nasal polyps
– Nasal inflammation:
– CT imaging
– Nasal polyp surgery:
– Need for nasal surgery
– Time to first nasal surgery
– Nasal surgery at 24 weeks
– Work productivity:
– WPAI-GH (Work Productivity and Activity Impairment — General Health)
– Harms outcomes:
– AEs (adverse events)
– SAEs (serious adverse events)
– WDAEs (withdrawal due to adverse events)
– Mortality
– Notable harms of special interest, including systemic and local injection site reactions; serious and opportunistic infections; serious cardiac, vascular, and thromboembolic events; and serious ischemic events.

**Study Designs**

– Published and unpublished phase III and IV RCTs (randomized controlled trials)

### SR0737-Kerendia

**Patient Population**

– Adults with chronic kidney disease and type 2 diabetes.
– Subgroups:
– Albuminuria at baseline
– eGFR (estimated glomerular filtration rate) at baseline
– SGLT2 (sodium-glucose cotransporter-2) inhibitor use at baseline
– History of cardiovascular disease
– Use of ACE (angiotensin-converting enzyme) inhibitor and/or ARB (angiotensin receptor blocker)

**Intervention**

– Finerenone 10 mg and 20 mg, oral administration.

**Comparators**

– Placebo plus SOC (standard of care, including an ACE inhibitor or ARB)
– SGLT2 inhibitor plus SOC

**Outcomes**

– Efficacy outcomes:
– Renal events (e.g., kidney failure)
– eGFR (estimated glomerular filtration rate)
– Urinary albumin-creatinine ratio
– Cardiovascular events (e.g., myocardial infarction)
– Mortality (renal, cardiovascular, and all-cause)
– Hospitalization (renal, cardiovascular, and all-cause)
– HRQoL (health-related quality of life)
– Symptom severity
– Functional status
– Harms outcomes:
– AEs (adverse events)
– SAEs (serious adverse events)
– WDAEs (withdrawal due to adverse events)
– Mortality
– Notable harms or harms of special interest (e.g., hyperkalemia, new onset of atrial fibrillation and atrial flutter, hypotension)

**Study Design**

– Published and unpublished phase III and IV RCTs (randomized controlled trials)

## Data Availability

All data produced in the present study are available upon reasonable request to the authors

## References

1. Cumpston M, Li T, Page MJ, et al. Updated guidance for trusted systematic reviews: a new edition of the Cochrane Handbook for Systematic Reviews of Interventions. The Cochrane database of systematic reviews. 2019;2019(10).

2. Borah R, Brown AW, Capers PL, Kaiser KA. Analysis of the time and workers needed to conduct systematic reviews of medical interventions using data from the PROSPERO registry. BMJ Open. 2017;7(2):e012545.

3. Michelson M, Reuter K. Corrigendum to “The significant cost of systematic reviews and meta-analyses: A call for greater involvement of machine learning to assess the promise of clinical trials” [Contemp. Clin. Trials Commun. 16 (2019) 100443]. Contemp Clin Trials Commun. 2019;16:100450.

4. Michelson M, Reuter K. The significant cost of systematic reviews and meta-analyses: A call for greater involvement of machine learning to assess the promise of clinical trials. Contemp Clin Trials Commun. 2019;16:100443.

5. Shemilt I, Khan N, Park S, Thomas J. Use of cost-effectiveness analysis to compare the efficiency of study identification methods in systematic reviews. Systematic Reviews. 2016;5(1):140.

6. Nussbaumer-Streit B, Ellen M, Klerings I, et al. Resource use during systematic review production varies widely: a scoping review. Journal of Clinical Epidemiology. 2021;139:287–296.

7. Kebede MM, Le Cornet C, Fortner RT. In-depth evaluation of machine learning methods for semi-automating article screening in a systematic review of mechanistic literature. Research Synthesis Methods. 2023;14(2):156–172.

8. Marshall IJ, Wallace BC. Toward systematic review automation: a practical guide to using machine learning tools in research synthesis. Systematic reviews. 2019;8:1–10.

9. De Bruin J, Ma Y, Ferdinands G, Teijema J, Van de Schoot R. SYNERGY - Open machine learning dataset on study selection in systematic reviews. V1 ed: DataverseNL; 2023.

10. Blaizot A, Veettil SK, Saidoung P, et al. Using artificial intelligence methods for systematic review in health sciences: A systematic review. Research Synthesis Methods. 2022;13(3):353–362.

11. van de Schoot R, de Bruin J, Schram R, et al. An open source machine learning framework for efficient and transparent systematic reviews. Nature Machine Intelligence. 2021;3(2):125–133.

12. Burgard T, Bittermann A. Reducing Literature Screening Workload With Machine Learning. Zeitschrift für Psychologie. 2023;231(1):3–15.

13. Khraisha Q, Put S, Kappenberg J, Warraitch A, Hadfield K. Can large language models replace humans in systematic reviews? Evaluating GPT-4’s efficacy in screening and extracting data from peer-reviewed and grey literature in multiple languages. Research Synthesis Methods. 2024;15(4):616–626.

14. Matsui K, Utsumi T, Aoki Y, Maruki T, Takeshima M, Takaesu Y. Human-Comparable Sensitivity of Large Language Models in Identifying Eligible Studies Through Title and Abstract Screening: 3-Layer Strategy Using GPT-3.5 and GPT-4 for Systematic Reviews. J Med Internet Res. 2024;26:e52758.

15. Landschaft A, Antweiler D, Mackay S, et al. Implementation and evaluation of an additional GPT-4-based reviewer in PRISMA-based medical systematic literature reviews. International Journal of Medical Informatics. 2024;189:105531.

16. Tran V-T, Gartlehner G, Yaacoub S, et al. Sensitivity and Specificity of Using GPT-3.5 Turbo Models for Title and Abstract Screening in Systematic Reviews and Meta-analyses. Annals of Internal Medicine. 2024;177(6):791–799.

17. Cohen JF, Korevaar DA, Altman DG, et al. STARD 2015 guidelines for reporting diagnostic accuracy studies: explanation and elaboration. BMJ Open. 2016;6(11):e012799.

18. CADTH Reimbursement Reviews and Recommendations. Darolutamide (Nubeqa): CADTH Reimbursement Review: Therapeutic area: Metastatic castration-sensitive prostate cancer. Ottawa (ON): Canadian Agency for Drugs and Technologies in Health; 2023.

19. CADTH Reimbursement Reviews and Recommendations. Durvalumab (Imfinzi): CADTH Reimbursement Review: Therapeutic area: Biliary tract cancer. Ottawa (ON): Canadian Agency for Drugs and Technologies in Health; 2023.

20. CADTH Reimbursement Reviews and Recommendations. Crisantaspase Recombinant (Rylaze): CADTH Reimbursement Review: Therapeutic area: Acute lymphoblastic leukemia. Ottawa (ON): Canadian Agency for Drugs and Technologies in Health; 2023.

21. CADTH Reimbursement Reviews and Recommendations. Upadacitinib (Rinvoq): CADTH Reimbursement Recommendation: Indication: For the treatment of adult patients with moderately to severely active ulcerative colitis who have demonstrated prior treatment failure, i.e., an inadequate response to, loss of response to, or intolerance to at least 1 of conventional and/or biologic therapy. Ottawa (ON): Canadian Agency for Drugs and Technologies in Health; 2023.

22. CADTH Reimbursement Reviews and Recommendations. Guselkumab (Tremfya): CADTH Reimbursement Review: Therapeutic Area: Psoriatic arthritis. Ottawa (ON): Canadian Agency for Drugs and Technologies in Health; 2023.

23. CADTH Reimbursement Reviews and Recommendations. Lumasiran (Oxlumo): CADTH Reimbursement Review: Therapeutic area: Primary hyperoxaluria type 1. Ottawa (ON): Canadian Agency for Drugs and Technologies in Health; 2023.

24. CADTH Reimbursement Reviews and Recommendations. Mepolizumab (Nucala): CADTH Reimbursement Review: Therapeutic area: Severe chronic rhinosinusitis with nasal polyps. Ottawa (ON): Canadian Agency for Drugs and Technologies in Health; 2023.

25. CADTH Reimbursement Reviews and Recommendations. Finerenone (Kerendia): CADTH Reimbursement Review: Therapeutic area: Chronic kidney disease. Ottawa (ON): Canadian Agency for Drugs and Technologies in Health; 2023.

26. Priem J, Piwowar H, Orr R. OpenAlex: A fully-open index of scholarly works, authors, venues, institutions, and concepts. arXiv preprint arXiv:220501833. 2022.

27. Van Rossum G, Drake FL. Python reference manual. Vol 111: Centrum voor Wiskunde en Informatica Amsterdam; 1995.

28. Pedregosa F, Varoquaux G, Gramfort A, et al. Scikit-learn: Machine learning in Python. the Journal of machine Learning research. 2011;12:2825–2830.

29. Reback J, McKinney W, Van Den Bossche J, et al. pandas-dev/pandas: Pandas 1.0. 5. Zenodo. 2020.

30. Harris CR, Millman KJ, Van Der Walt SJ, et al. Array programming with NumPy. Nature. 2020;585(7825):357–362.

